# Automated Airways Characterization and Assessment of Cystic Fibrosis from CT Imaging

**DOI:** 10.64898/2026.06.09.26355170

**Authors:** Juan Antonio K. Chong Chie, Matthew L. Cooper, Scott A. Persohn, Charles P. Burton, Paul Salama, Paul R. Territo

## Abstract

**Background:** Advancements in medical imaging have enabled non-invasive diagnosis and staging of cystic fibrosis (CF) using CT scans, revealing dilated airways, an increased number of visible airways, and airway generation splits in these patients. However, manual characterization of airways remains time-consuming and challenging due to the numerous structural changes, thereby limiting clinical feasibility. This study aims to develop an automated algorithm to characterize airways from segmented lung CT scans and apply this to a retrospective population. This approach reduces the time required to analyze images and obtain disease-staging results.

**Methods:** This framework consists of two stages. The first stage extracts and skeletonizes the airway tree from lung CTs, while the second stage measures lung features, including airway volumes, branch counts, generation splits, diameters, and cross-sectional areas. This permits comprehensive characterization for use in clinical assessment.

**Results:** The airways analysis was performed on 169 CT volumes ranging in age from 6 to 18 years of age, revealing substantial differences in detected airway branches, generation splits, and normalized airway volume between the control and CF groups. The framework also measures airway diameters and cross-sectional areas, revealing an increase in the number of small airways in cystic fibrosis patients, due to early bronchiectasis. These findings align with previous research and demonstrate the framework’s ability to accurately quantify airway changes in patients with CF.

**Discussion:** The framework extracts entire airway trees, facilitating measurements of volume, branch count, diameters, and cross-sectional areas, which change with CF severity and/or treatment. However, partial lung atelectasis can limit the accuracy of airway detection in moderate-to-severe cases.

**Funding:** NIA U54 AG054345 and NIA R21 AG07857501

## 1. Introduction

Cystic fibrosis (CF) is a life-shortening genetic disorder resulting from mutations in the cystic fibrosis transmembrane conductance regulator (CFTR) gene. The disease is associated with an average lifetime cost of $306,322 (USD) per patient^1–3^, with an incidence of approximately 1 in every 3,400 births,^4,5^ and leads to an estimated global disease population of 162,428 annually.^4,5^ The CFTR gene encodes a protein responsible for maintaining osmotic balance in the body by transporting chloride ions across cell membranes, which regulates water movement.^1,6^ Proper function of this protein ensures that mucus remains thin, enabling airway cilia to clear mucus and debris from the lungs efficiently.^1,6^ However, CFTR gene defects result in insufficient or absent CFTR protein production results in chloride ion retention within cells and impaired water transport.^7^ As a result, this dysfunction causes mucus to accumulate as a thick and concentrated mass that the cilia cannot expel.^7^ Over time, excessive production of viscous mucus leads to airway obstruction, reduced pulmonary function, increased cellular infiltration, and persistent lung infections.^5,6^ Diagnosis of CF typically involves a multimodal approach, including a sweat chloride test, genetic testing, and a clinical evaluation longitudinally by an accredited care center.^6,8^

Advancements in medical imaging have enabled non-invasive diagnosis and staging of CF through high-resolution computed tomography (CT) imaging.^9^ CF patients commonly present with lung abnormalities (e.g., bronchiectasis, atelectasis due to mucus plugging, and parenchymal opacities) that alter lung anatomy.^10,11^ These pathological changes reduce the accuracy of classical segmentation techniques (e.g., threshold-based, shape-based)^12^ and compromise staging scores due to the temporal and spatial variability in disease distribution and extent.^13^ Studies have also shown that CF patients display increased numbers of visible and dilated airways, as well as more detectable generation splits on CT scans.^14,15^ These factors complicate lung extraction and segmentation in the context of CF-related pathology.

Although airway segmentation remains challenging, recent studies show that quantifying airway abnormalities (e.g., number of visible airways, average airway diameter) can characterize structural lung disease in CF and may help assess disease severity, early progression, and response to therapy, providing valuable insights into the extent and severity of CF.^14,15^ Currently, airways are manually assessed for abnormal diameters and visible branchings, requiring experts to locate, evaluate, segment, and measure each airway slice-by-slice.^8,16^ This approach is time-intensive and requires specialized anatomical and clinical knowledge, limiting its practicality for routine clinical use.^16^ Importantly, this method does not account for additional lung lesions or disease manifestations, which are clinically recognized as surrogate markers of disease stage.^15^

The primary challenges of manual analysis include the large number of airways, structural alterations in airway morphology due to pathology, the extensive number of slices in high-resolution CT volumes, and the difficulty in distinguishing obstructed airways from blood vessels.^17^ Consequently, manual airway characterization is neither practical nor feasible for routine clinical application.^18^

Clinical pulmonary function testing (PFT) is widely regarded as the gold standard for assessing lung function in children aged six years or older and in adults,^19^ and it is routinely used to evaluate lung function in individuals with CF for clinical management.^1,19^ These PFT metrics reflect the integrated function of the lung, which is influenced by clinical manifestations (e.g., airway wall thickening, parenchymal disease, mucus plugs);^20,21^ however, previous studies have demonstrated that PFT lacks sensitivity for detecting subtle changes.^22,23^ In contrast, non-contrast CT imaging has been shown to be more effective and sensitive for identifying structural lung damage that are linked to functional PFT manifestations.^2^ In addition, CT imaging is more feasible for monitoring lung changes in patients unable to perform spirometry, including neonates, infants, toddlers, and preschoolers.^3^

This study aims to develop, implement, and validate an automated framework for accurate segmentation of the lungs and trachea, and for quantitative airway characterization from CT scans of CF patients. In addition, given the limitations of PFT, this study aims to elucidate the relationships between functional measures and airways characterization. The automated framework reveals significant differences in disease-related metrics between control and CF cases, consistent with previous studies.

## 2. Methods and Materials

### 2.1 Dataset

The dataset used consists of 169 CT scans collected between 2012 and 2021 by IU Health across 7 clinical sites. Subjects were aged 6 to 18 years, with a sex distribution of 51% male and 49% female. Among these, 49 were healthy controls (59.2% male, 40.8% female), and 120 were CF patients (47.5% male, 52.5% female) spanning a range of severity stages (mild = 51, moderate = 36, severe = 33), as determined by the Brody^24^ scoring system. All images were provided by IU Health and de-identified in accordance with HIPAA regulations. Detailed demographic statistics and imaging properties are presented in Figure 1A. In addition, spirometry measurements for forced vital capacity (FVC), forced expiratory volume in one second (FEV1), and peak expiratory flow rate (PEFR) were obtained for these subjects. A subset of 18 control cases and 54 CF patients, with varying severity stages, was manually segmented by three experts. This dataset of 72 segmented lungs was used to evaluate the performance of the lung and trachea segmentation algorithm.

**Figure 1.**
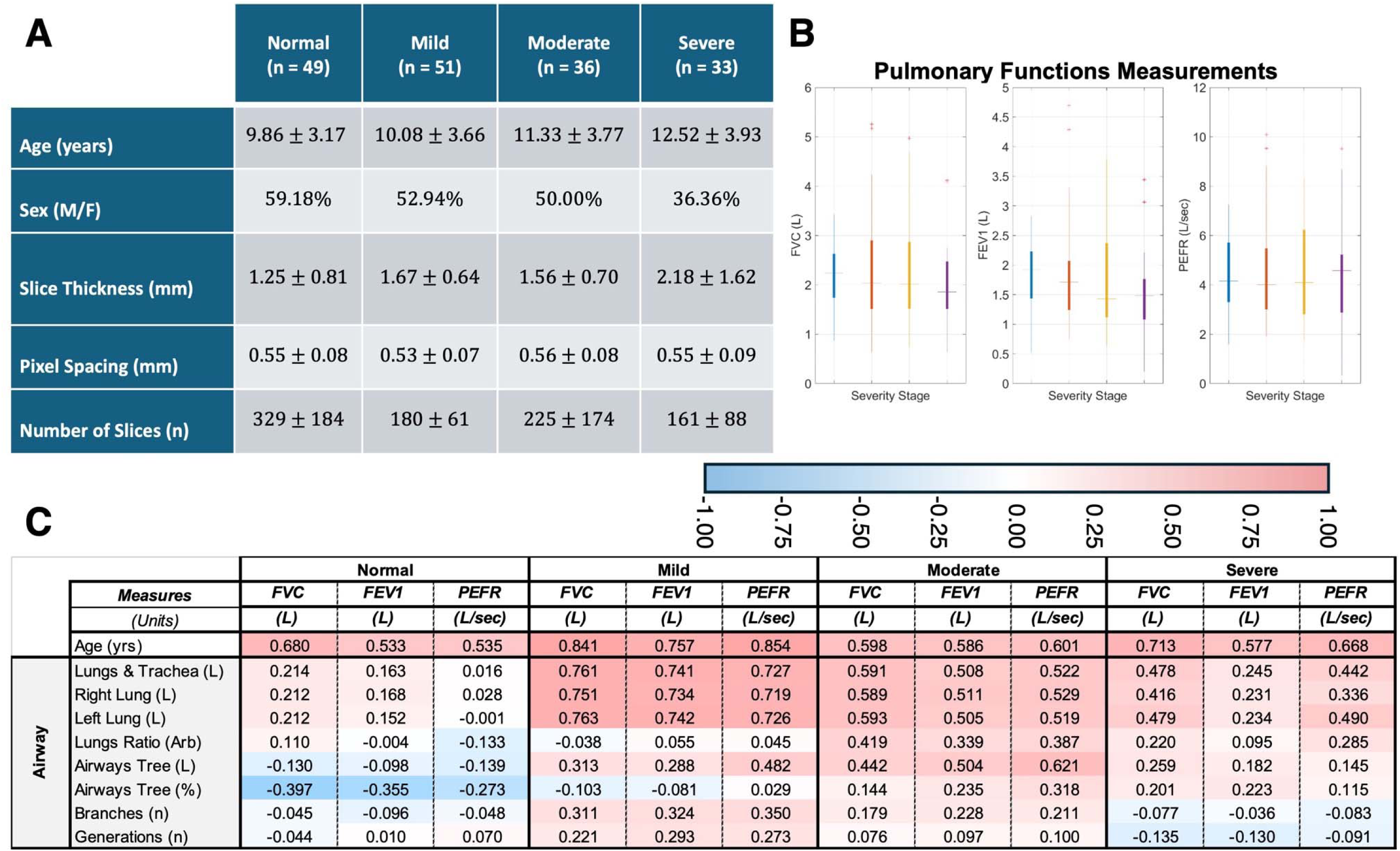
Subject Demographics, Pulmonary Function Tests, and Correlations with Pulmonary Measurements. (A) Demographics of the Dataset. This information was extracted from a dataset consisting of 169 CT scans, of which 49 are from control patients and 120 from CF patients. Data are presented as Mean±SD. (B) Pulmonary function test (PFT) box-and-whisker plots across the CF spectrum, where the tests did not show significant differences across disease stages. (C) Pearson Correlations of airways characteristic measurements with PFT, which include forced vital capacity (FVC), forced expiratory volume in 1 second (FEV1), and peak expiratory flow rate (PEFR). Data are color-coded according to the correlation value over the interval [-1.0,1.0].

### 2.2 Lung and Trachea Segmentation

The segmentation process consists of two parallel methods: extraction of the trachea from a CT volume and segmentation of the lungs (Figure 2A). Trachea segmentation in normal subjects can be relatively straightforward; although CF is associated with tracheal abnormalities in some patients making segmentation more challening.^25–27^ To overcome this, a 3D region-growing approach is used, leveraging the similarity between tracheal voxel intensities (in Hounsfield Units; HU) and the HU value of air (−1000 HU). The algorithm does not require iterating over the entire CT volume because the trachea is typically present only in the initial 50% to 60% of slices; thus, iterating over the entire volume results in unnecessary resource expenditure. To optimize computational efficiency, the algorithm processes the first 65% of slices (as a safety margin), starting from the sub-clavicular notch and extending toward the lung base.

**Figure 2.**
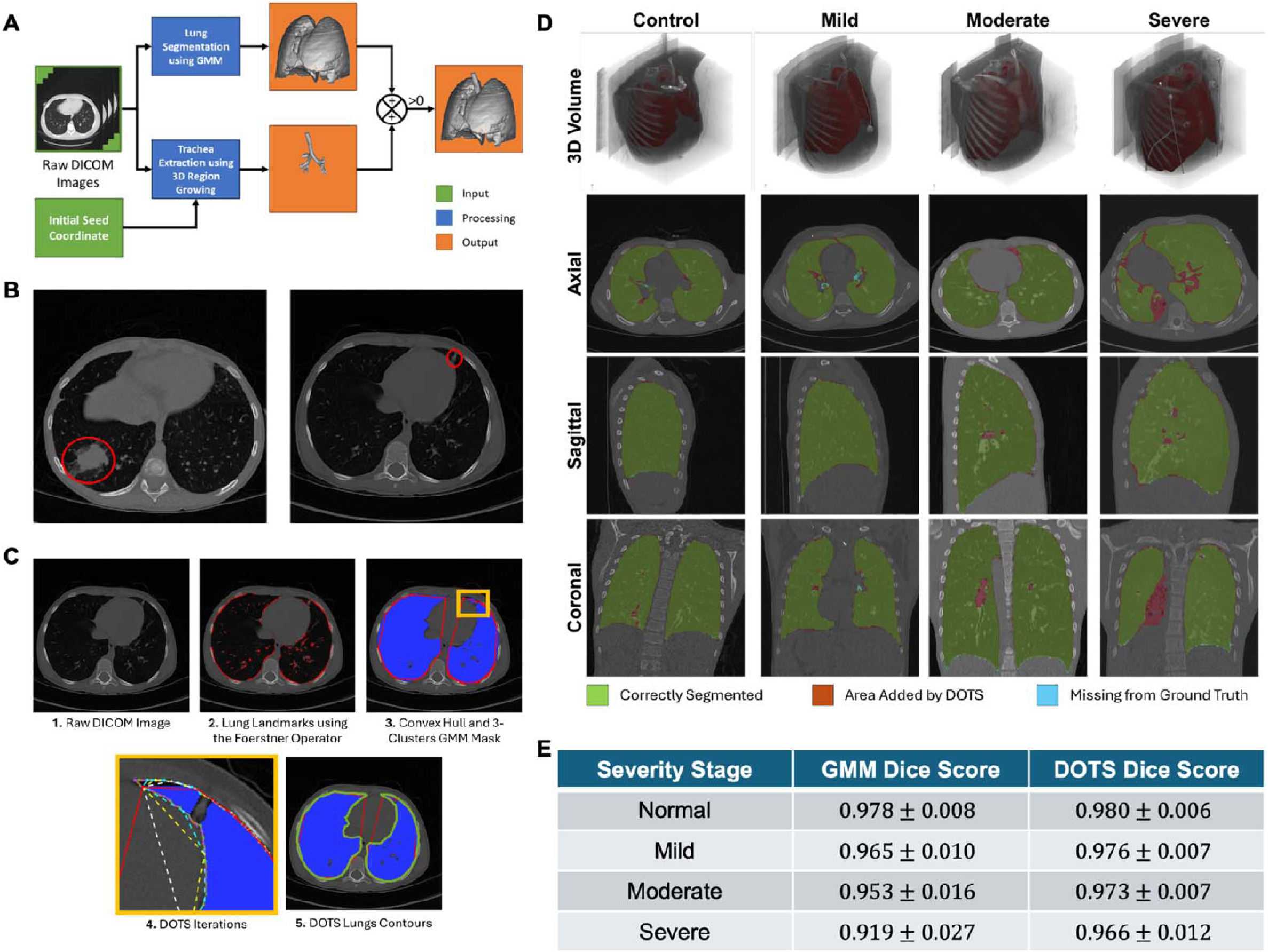
DOTS Algorithm and Segmentation Results. (A) Graphical depiction of the proposed lung and trachea segmentation framework. (B) Example of lungs with consolidated regions (red). (C) Graphical depiction of the steps of the DOTS algorithm. (D) Example of segmented lung volumes using the proposed framework for different CF severity stages. The Dice scores for each case are: Control = 0.9842, Mild = 0.9790, Moderate = 0.9713, and Severe = 0.9615. (E) Average Dice Score by CF severity stage.

In contrast, lung segmentation in CF patients poses numerous challenges due to pathological abnormalities (Figure 2B). In CF staging, images were acquired during inspiration, which can cause the anterior and posterior lung junctions to appear fused in some cases. Therefore, the combination of lung anatomy, non-uniform lesion distribution, and ill-defined structural boundaries further complicates segmentation. To address these challenges, a multi-stage lung segmentation pipeline was developed, which uses a three-cluster Gaussian mixture model (GMM),^28^ a 26-neighbor connected components model,^29^ and, when the GMM fails to delineate the entire lung, a novel iterative algorithm called Delineation of Thoracic Structures (DOTS) (Figure 2C).

Lung segmentation begins by computing the histogram of HU values for voxels in the current slice, accounting for structural shape changes at each slice location within the volume. For instance, the lung appears quasi-circular at the top and transitions to a thin “C” shape at the base (Figure S1A-C). Consequently, the HU histogram and peak locations vary with slice position (Figure S1D-F). Hence, to account for these location-dependent shape changes, the histogram is constructed using the current slice and its adjacent slices (previous and next), rather than relying on a single slice alone.

After constructing the histogram, a three-cluster GMM is fitted under specific conditions: (1) the first cluster peak must be within [−1000, −900] HU; 2) the third cluster peak must be within [0, 1000] HU; (3) the second cluster peak must be located between the first and third cluster; (4) each cluster peak the other two clusters (i.e. 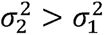 and 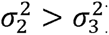)). These constraints ensure that the first cluster value must be unique; and (5) the variance of the second cluster must be greater than the variance of centers around the HU value of air, the second on the transitional region between air and soft tissue (where voxels are a mix between air and soft tissue, having a flatter distribution), and the third around the HU value of soft tissue and muscle (Figure S2A-C). Due to structural changes across slices, histograms and cluster intersection cutoff points vary by slice. The cutoff points 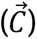 between clusters *i* and *j* for slice *k* are determined by Equation 1:

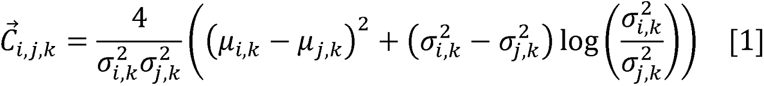

where *i* and *j* are the clusters, *k* is the slice index, *μ_i_* is the mean of the *i*-th cluster, and *σ_i_* is the standard deviation of the *i*-th cluster. Each voxel is then classified into its respective GMM cluster based on its HU value and spatial alignment. Following initial classification, segmentation masks are refined by evaluating a 3×3×3 neighborhood around each voxel; the cluster with the highest voxel count in this neighborhood is assigned to the central voxel.

In severe CF cases, patients may exhibit partial lung collapse due to atelectasis, pleural effusions, or lung parenchymal diseases (Figure S2D-F). These conditions manifest as fissures or consolidated regions with HU values closer to soft tissue ([13, 300]). Because atelectasis, pleural effusion, and parenchymal opacities are CF-related lesions relevant for disease staging,^10,11^ regions affected by these abnormalities must be included in the segmented lung mask. However, the HU values of these abnormal voxels often fall outside the lung tissue cluster, leading to potential misclassification as body voxels. To overcome this limitation, DOTS algorithm was developed for cases where the GMM fails to segment the lungs due to atelectasis, pleural effusions, or lung parenchymal diseases. The method begins by extracting the body wall contour on each CT slice, followed by the detection of lung landmarks within this contour using the Foerstner operator.^30^ Next, each lung landmark is classified as left or right, and a coarse convex hull^31^ is constructed for each lung using these landmarks. Then, for each convex hull, DOTS refines the segmentation by iteratively selecting vertex pairs on the convex hull, constructing midpoints, and generating orthogonal projections to the lung wall from each midpoint (Figure 2C). New vertices are added to the contour, and the process repeats until the distance between adjacent vertices matches the image resolution, yielding a final contour for each lung per slice. A limitation occurs when the algorithm encounters Foerstner landmarks that cannot be correctly classified as left or right, resulting in both lungs being clustered into a single convex hull. Figure 2C shows a graphical depiction of DOTS for a trans-axial slice, and the corresponding pseudocode is provided in Algorithm 1.

#### Algorithm 1 DOTS Algorithm

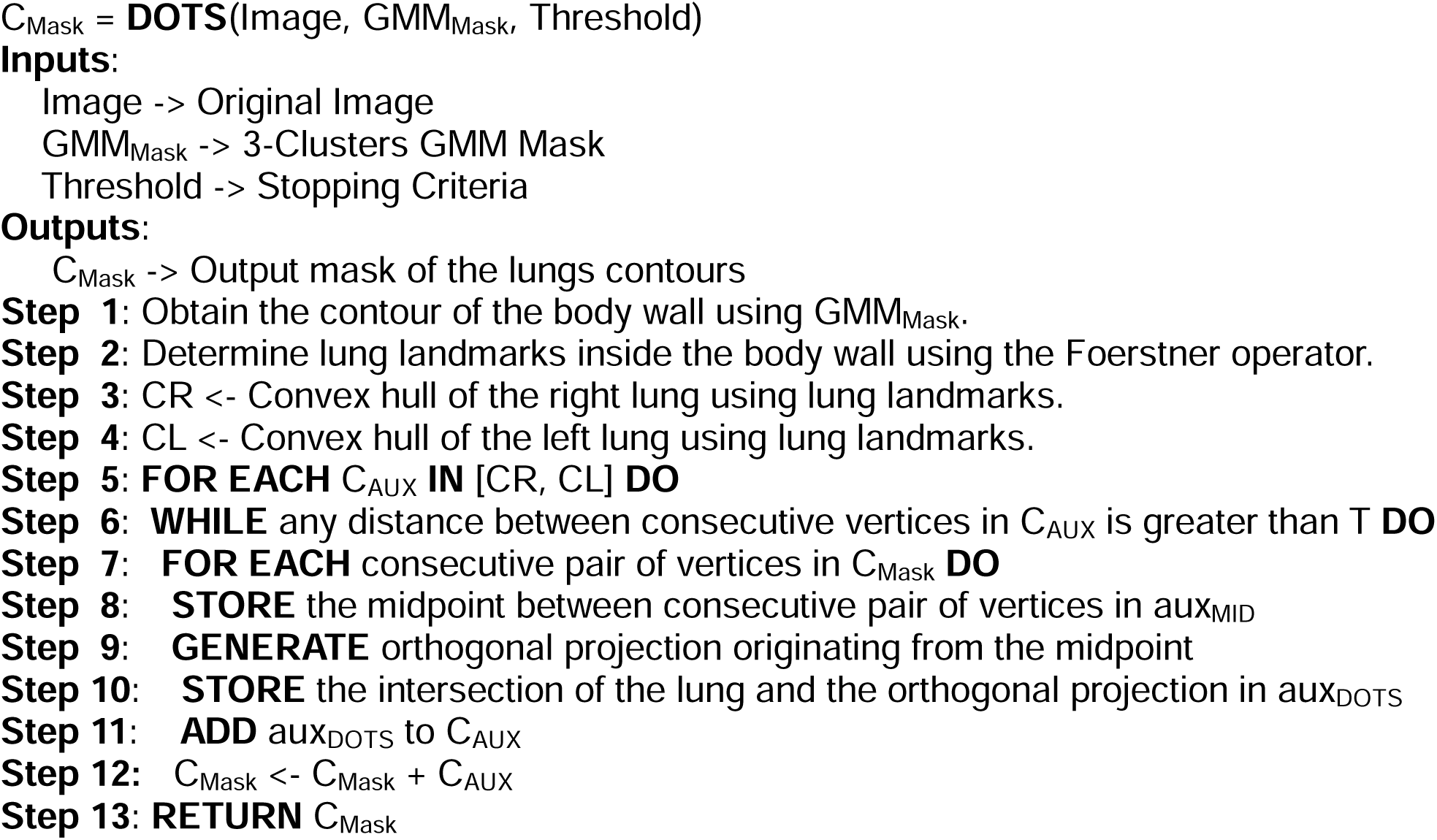

To ensure complete lung separation, a further refinement step to DOTS was added, where an iterative approach using morphological erosion and Euclidean distances, called Separation of Lung Volume Regions (SOLVR; Figure S3A), was developed. This method identifies and processes only those slices where the lungs appear fused (Figure S3B), often due to anatomical structures (e.g., anterior sterno-pericardial ligaments; Figure S3B inset). The process starts by removing the trachea from the binary lung and trachea masks via element-wise mask multiplication. Then, the algorithm identifies and counts connected components, evaluating the success of lung separation according to the following rules:

- The number of 3D connected components created is two or greater.
- If there are more than two connected components, the number of voxels comprising the two largest components must be at least 90% of the total number of voxels.
- The difference in the number of voxels between the two largest connected components must be less than 10% of the total number of voxels.
- The location of the centroids of the largest components cannot be in the same half of the image along the x-axis.
- The overlap between the bounding boxes of the two largest components cannot be more than 20%.

If at least three of the five criteria are satisfied, separation of the left and right lungs is deemed successful, and the process stops. Otherwise, the lungs are considered connected, and a secondary 2D iterative approach is used. This method identifies the specific slices where fusion occurs (two or less criteria are satisfied), and performs separation only on those slices. For each identified slice, iterative 2D erosion is conducted until the lungs are separated. The same set of rules, with minor adjustments to scale down from 3D to 2D, is used to assess separation. For instance, if, after a set number of iterations, the lungs remain connected, the original slice is down-sampled by a factor of two, and the erosion process is repeated. During testing and deployment, it was found that the algorithm did not require more than four iterations of the erosion process before the down-sampling step. Upon successful separation, the algorithm extracts the region of connection, defined as the area of eroded voxels between separated lung volumes. If down-sampling was performed in the previous step, this region is up-sampled to the original resolution. Next, for each eroded voxel, the shortest Euclidean distance to the perimeter of both lungs is computed, yielding two distance maps indicating the proximity of the eroded voxel to each lung boundary. Lastly, these maps are averaged to create a combined distance representation, with smaller values indicating closer proximity to both lungs and balancing the contributions from both sides. The voxel with the smallest value in this average map is designated as the voxel of connection, representing the most probable boundary point for separating the left and right lung. Figure S3C shows the results for a single slice, and the pseudocode is detailed in Algorithm 2.

#### Algorithm 2 SOLVR Algorithm

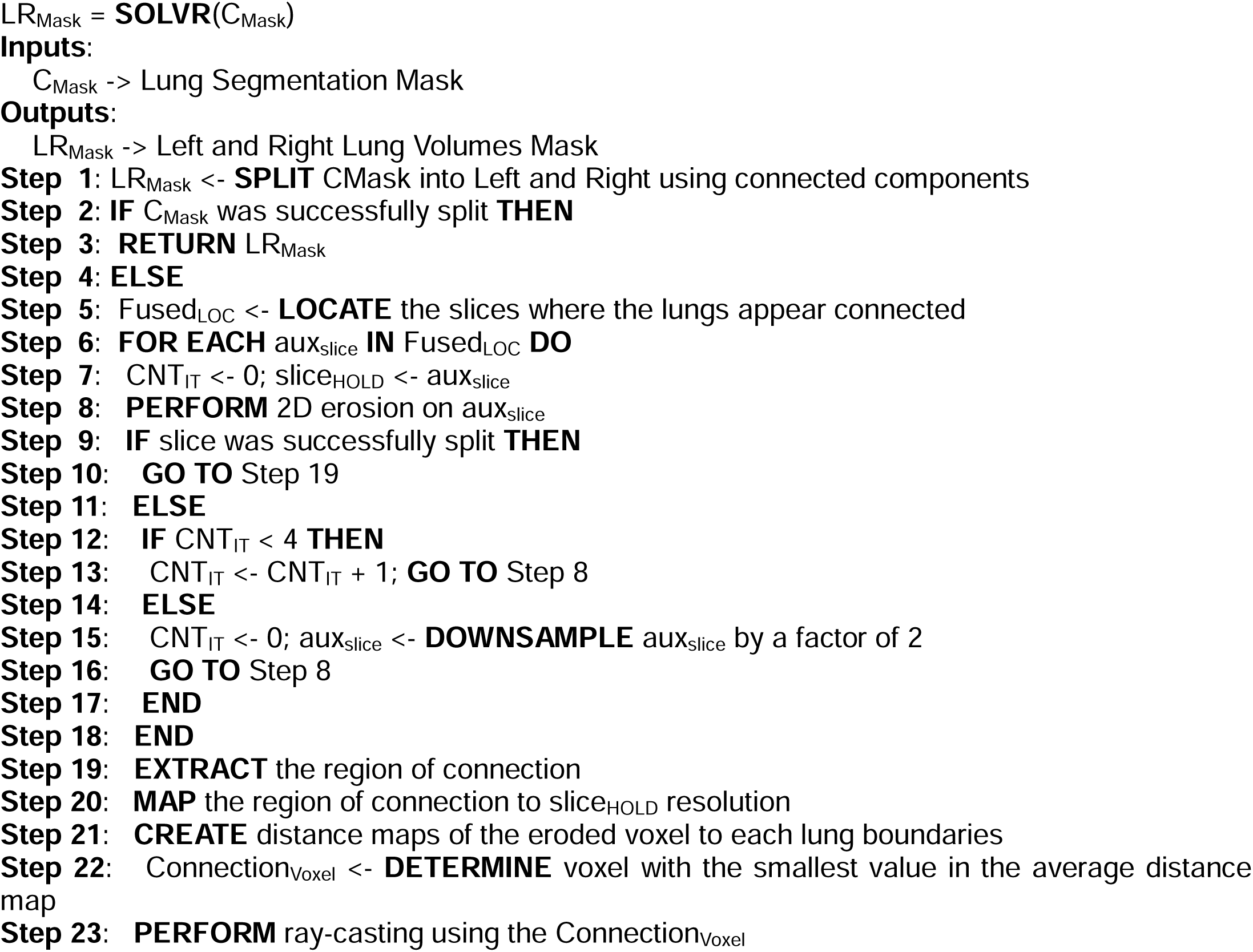
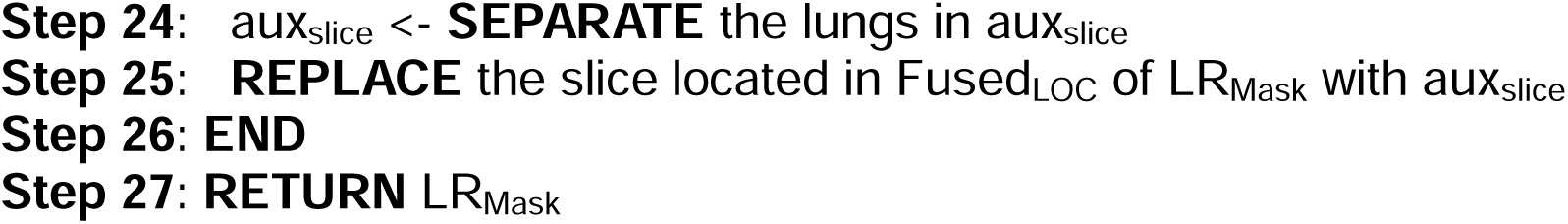

To determine the separation planes’ orientation, a ray-casting procedure projects eight rays from the correction voxel at 45° intervals, extending them until they reach background voxels outside the lung. The pair of non-consecutive rays with the shortest distance to background voxels defines the separation plane. This plane eliminates voxels along each selected ray from the correction voxel to the background voxel.

### 2.3 Airways Analysis

Automatic identification of potential airway voxels is achieved using a two-stage framework (Figure 3A). By modeling the histogram of HU values for segmented lung voxels as a two-cluster GMM (Figure 3B), the first stage uses particle swam optimization^32^ to find the optimal values that determine the HU interval for the airway wall. The second stage applies thresholding based on these intervals to obtain candidate airway voxels. Airway wall voxels, which have intensities higher than lung tissue and air but lower than those of blood vessels and fibrotic tissue, are classified into the rightmost cluster (Figure 3B). To differentiate between airway and blood vessel voxels, the airway lumen is systematically traversed from the trachea to the diaphragm, permitting exclusion of vascular voxels. Subsequently, airways are extracted from candidate voxels using a 26-neighbor connected-component model,^29^ starting at the top of the trachea. Figure 3C shows the segmented lung mask used for airways analysis, and Figure 3D shows the extracted airway tree.

**Figure 3.**
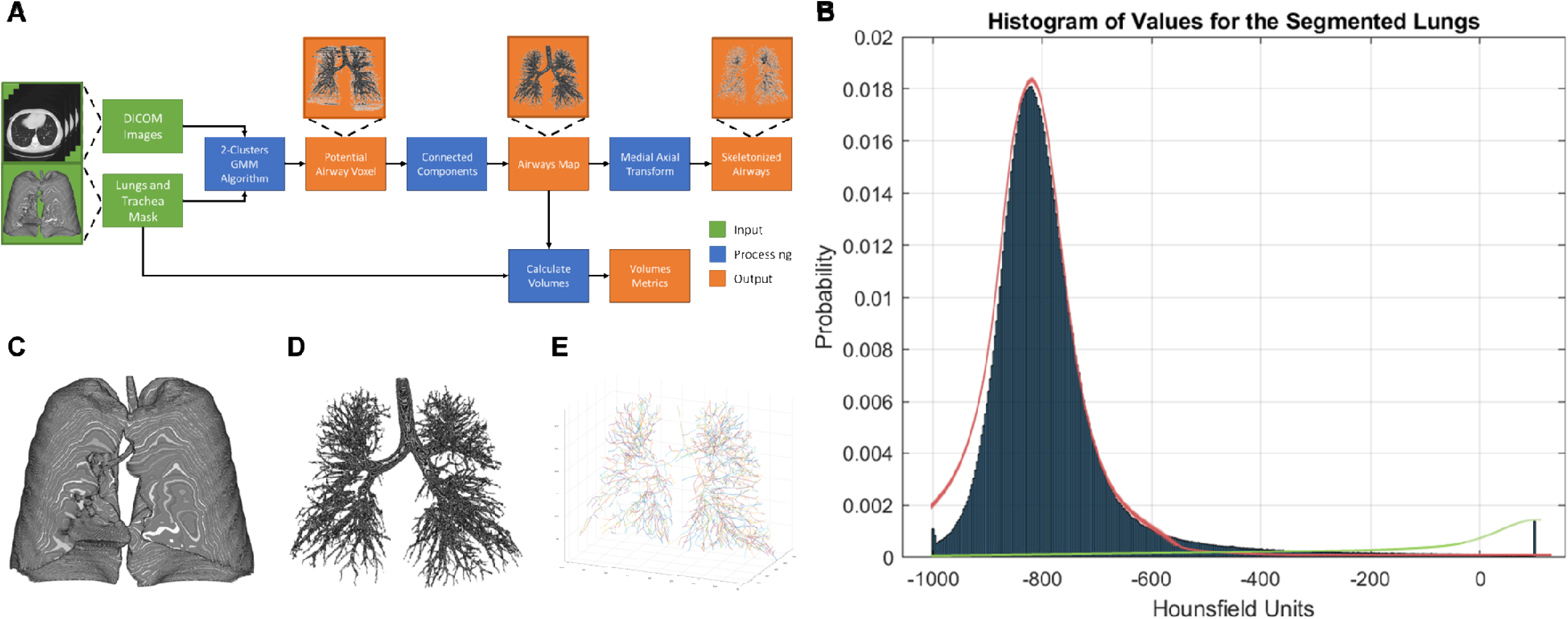
Airway Characterization Framework. (A) Graphical depiction of the airway extraction, airway skeletonization, and volumes measurement stage. (B) Histogram of Hounsfield units (HU) values of the segmented lung volumes for a single case. 3D views of the obtained (C) segmented lung mask, (D) extracted airways tree, and (E) skeletonized airways tree.

After extracting the airway tree, the airway lumen is filled, and skeletonization performed using the medial-axis transform.^33^ This process applies successive morphological erosion to thin the structure while preserving endpoints, ultimately generating a midline skeleton of the original airway tree (Figure 3E). The skeletonized airway tree facilitates counting branches and identifying generation splits. The center of the airway is estimated following extraction and skeletonization. Figure 3E provides an example of the skeletonized airway tree produced by this method.

Airway characterization is performed by obtaining the following metrics for the whole airway tree: lung volumes, volume of the airway tree, percentage of the lungs occupied by the airway tree, number of detected airways branches, and number of detected generation splits (Figure 4A). To count the number of branches, the framework starts by finding the locations where the airways split in the skeletonized airway tree. This task is performed by traversing the skeletonized airway, starting with the top of the trachea, following the path of connected voxels, and enumerating each voxel where the path divides (Figure 4B). A branch is then defined as all the connected voxels between two split locations. To quantify the generations, the framework labels each branch based on the number of times a given path is divided, with the trachea being the first generation. For example, when the trachea divides into the primary bronchi, these two branches representing the bronchi will be labeled as first generation.

**Figure 4.**
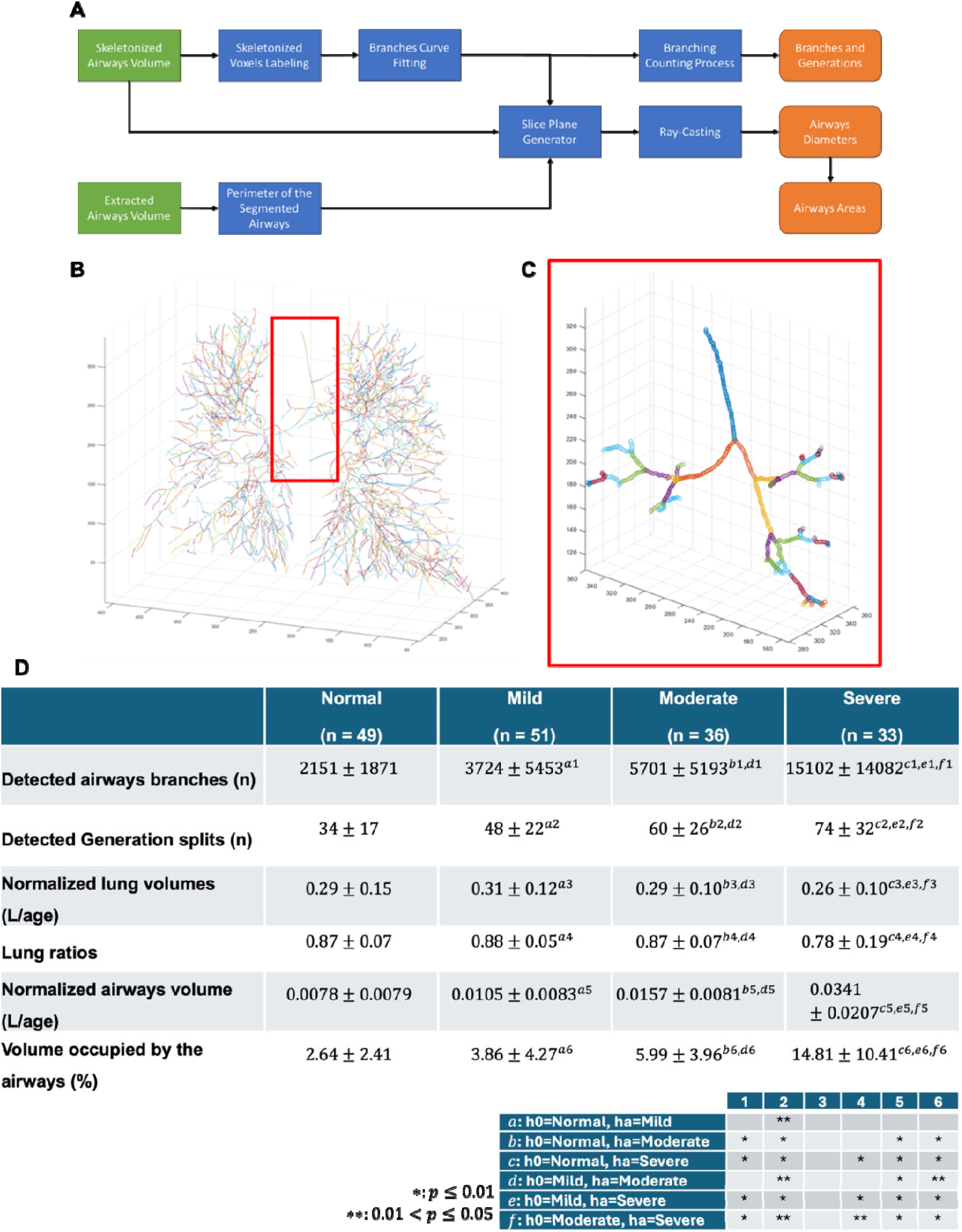
Airway Characterization and Results. (A) Graphical depiction of the airway characterization stage. (B) Results of the quantification of airways branches and generation splits in the complete airway tree. (C) Zoom-in of the trachea area from B. (D) Quantitative averages measures of lung volume, lung volume ratio, airway tree, number of branches, and segmental generation as a function of clinical diagnostic stage. Data are presented as Mean±SD

As the diameters of airway branches decrease with each division, the smallest branches can be as small as 0.51 mm,^34^ depending on the in-plane scanner resolution, reconstruction algorithm, and patient age. Cross-sectional areas of an airway can be modeled as an ellipse,^35^ and the orthogonal diameters of the ellipse are clinically reported. Hence, for whole-airway characterization, diameters and cross-sectional areas are measured branch-wise, approximating each branch as a flexible cylindrical structure orthogonal to the airway’s center axis (Figure 5A). These measurements are reported as histograms of airway diameters and cross-sectional areas.

**Figure 5.**
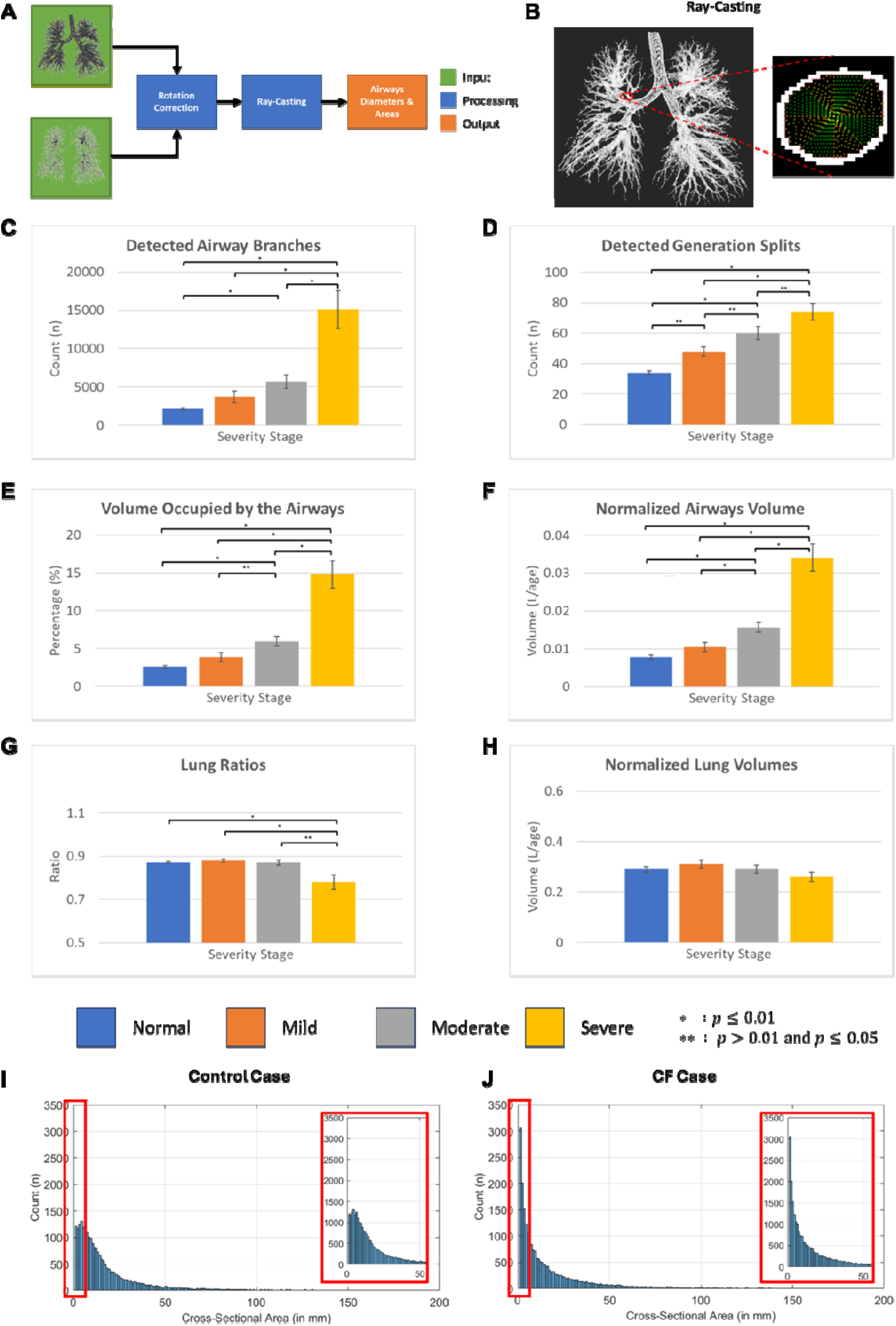
Ray-Casting Process and Results Comparison Across Disease Stage. (A) Graphical depiction of the ray-casting process. (B) Visualization of the ray-casting process at one location. Quantitative measures of (C) detected airways branches, (D) detected generation splits, (E) volume occupied by the airways, (F) normalized airways volumes, (G) lung ratios, (H) normalized lung volumes as a function of clinical diagnostic stage. Frequency analysis of airway cross-sectional areas as a function of clinical diagnosis for (I) control case and (J) CF case.

To perform this process, the framework fits a curve to the voxels that form each branch to approximate its center. A normal vector between consecutive voxels is calculated, and an orthogonal plane is placed midway between the voxels. Then, a 2D slice projection of the airway tree is generated using the orthogonal plane, the fitted curve voxel location, and the normal vector (Figure 5B). Provided this, a ray casting process with two perpendicular rays is performed from the fitted curve voxel, recording the intersection points with the airway contour on each side (Figure 5B inset), and the airway diameter is given by the distance between the two intersections. This ray-casting process is repeated by rotating the rays from 0 to 90 degrees in 1-degree increments to cover all possible rotation angles. The measured airway diameter is computed from the pair of diameters that yield the largest ellipse area using a prolate ellipsoid model.

### 2.4 Statistical Analysis

In this study, the lung ratio is defined as the minimum volume between both lungs divided by the maximum volume of the two lungs. This metric ranges from 0 to 1, where 0 indicates a complete absence of one lung and 1 indicates perfect symmetry. The lung ratio quantifies the degree of lung-volume asymmetry and may indicate disease progression or structural abnormalities.

Because CF is a genetic disease, the severity of lung involvement is generally determined by underlying gene mutations rather than patient age.^36^ However, lung volumes vary with individual body characteristics (e.g., height, weight, body mass index), which change with age.^37^ To minimize the impact of age-related body size differences on volumetric metrics and better isolate disease-specific effects, all volumetric measurements were normalized by patient age. The resulting age-normalized lung ratio allows for more meaningful comparisons across patients and disease stages.

All measured metrics (e.g., lung ratios, number of visible airways, percentage of the lung volume occupied by the airway tree) were analyzed using one-way ANOVA. Statistical significance was defined as p<0.05 between groups. In addition, correlation between all measured metrics and pulmonary function tests metrics were computed via Pearson’s correlations coefficients.

## 3. Results

### 3.1 Lung and Trachea Segmentation

Lung and trachea segmentation methods were evaluated using 72 CT scans. These included 18 control cases and 54 cases of CF at various stages of severity. Three trained personnel generated ground-truth volumes. The segmentation methods were compared after both the GMM and DOTS segmentation stages. Figure 2D presents segmented lung volumes across each of the four stages (control, mild, moderate, severe).

Segmentation performance was assessed using the Dice score,^38^ a metric that measures overlap between objects, which in this case are the two-stage framework results and the ground-truth. Overall, the two-stage segmentation method achieved an average Dice score of 0.974±0.010 across all volumes and severity cases. Notably, the largest benefit was observed in severe cases, where the Dice score increased from 0.919±0.027 at the GMM segmentation step to 0.966±0.012 at the DOTS refinement step, representing a 5.11% increase. In comparison, controls and mild cases showed much smaller performance gains (2% for controls and 1.14% for mild) between both stages, highlighting the pronounced improvement in the most challenging cases. For results on individual disease stages, see Figure 2E.

### 3.2 Airways Analysis

The airways analysis stage was conducted on 169 CT scans. Of these, 120 were from CT patients with disease stages ranging from mild to severe, as determined by a board-certified radiologist. There were also 49 scans from healthy control subjects. Lung volumes and corresponding airway trees were obtained and visually checked for quality assurance of each scan. The segmented lung mask was used to extract the lungs from the DICOM volume, enabling subsequent analysis to be focused on the lung region. Then, airway trees were skeletonized using the medial-axis transform. Visual examples of these volumes are shown in Figures 3C-E.

The skeletonized airway trees enabled quantification of the number of detected branches and generation splits. Figure 4B shows a visual representation of the characterized skeletonized tree. Each color denotes a distinct branch, defined as all connected voxels between two split locations. To facilitate visualization, Figure 4C presents a zoom-in view of the trachea. Figure 4D summarizes the number of detected branches, generation splits, normalized lung volumes, normalized lung ratios, and normalized airway volumes for each disease stage. Their statistical significance is also shown.

The results show significant differences in measurements between control and CF subjects. Specifically, the number of detected branches and generation splits is substantially higher in CF patients than in controls, consistent with previous reports.^14,15^ Similarly, normalized airway volumes also increase with disease progression. These significant findings (Figure 5C-G) exhibit an upward trend with increasing disease severity. In contrast, no significant changes were observed in the normalized lung volume metric (Figure 5H).

The airways analysis framework also measures the outer diameters of detected airways and calculates their cross-sectional areas, approximating them as elliptical shapes. Figures 5I-J present histograms of airway areas for a control subject and a CF patient of the same age with similar lung volumes. The results show that CF patients have a greater number of small airways detected than controls. This finding aligns with previous reports,^39,40^ which attribute increased detectability to irreversible damage from bronchiectasis or mucus plugging in the small airways. These findings are consistent with the trends observed in the number of detected branches and normalized airway volumes shown in Figure 4D.

Pearson correlations were computed to examine the relationship among PFT, airway metrics, and disease stage. However, PFT measures (FVC, FEV1, PEFR) did not significantly differ across clinical CF stages (Figure 1B). In contrast, the airway measured metrics exhibited a positive relationship with disease progression and were significantly different across and between stages (Figures 4D and 5C-H). Consistent with the PFT data across disease stages (Figure 1C), correlations with image-derived airway metrics showed a trend from negative to positive associations with airway tree volume, branch numbers, and the number of segmental generations.

## 4. Discussion

As previously described, the increase in the number of visible airways and the generation splits is attributed to lung lesions in the airways caused by conditions associated with CF, such as bronchiectasis and bronchial wall thickening.^10,11^ Upon analyzing data from mild and moderate cases, this irreversible damage to the airways manifests in CT scans as an increase in the area and volume occupied by the airway walls (Figures 4D and 5E), a change in the diameter of the airway branch (Figures 5I and 5J), or a combination of both. Building on these findings, we observed that airway branches and generation splits are significantly larger in individuals with CF compared to healthy controls (Figures 4D, 5I, and 5J). Based on this observation, we investigated the relationship between airway branching patterns and the clinical stage of the disease. Our framework automatically identifies airway split locations from the skeletonized tree, enabling the quantification of the number of branches as a function of CF severity. Notably, the analysis revealed pronounced stage-dependent changes, with increasing airway branching complexity correlating with advancing CF severity. In contrast, comparison of PFT measures with disease progression showed no significant changes (Figure 1B). Furthermore, correlations between PFT and airway metrics shifted from negative to positive, with the strongest associations observed at the mild stage. This finding suggests that airway characterization is particularly useful for assessing early disease remodeling of the airway. These results are consistent with established CF CT scoring systems (e.g., Brody, PRAGMA-CF^41^), which quantify the extent and severity of bronchiectasis, airway wall thickening, mucus plugging, and air trapping.^24,42^ Therefore, this approach offers a complementary descriptor of airway-tree organization by directly measuring visible branches and generation splits.

In addition, the airways analysis framework reveals a significant increase in the volume occupied by detected airways, as well as the number of detected airway branches and generation splits as the severity of CF progresses (Figures 4D, 5C, 5D, and 5F). In contrast, lung ratios and measured lung volumes remain relatively stable as disease stage severity increases (Figures 5G and 5H). These findings suggest that airway characterization may provide a sensitive quantitative biomarker for assessing CF progression and disease severity, complementing prior work showing that CT-based airway metrics track structural lung disease and may detect changes not captured by conventional measures.^14,15^ The increases in airway volume and branch count could be attributed to the presence of CF-related lung lesions, such as small airway mucous plugging that manifests as tree-in-bud nodules, and bronchiectasis, which make small airways more detectable and visible via CT. Accordingly, CF patients may exhibit a greater number of detectable small airways, likely because wall thickening and mucus plugging can enlarge or outline peripheral airways that are otherwise smaller than the scanner’s resolution, thereby increasing their apparent caliber and visibility on imaging.

Although the proposed framework shows an increasing trend across CF stages, its performance may be limited in advanced disease stages, where airway extraction becomes more difficult because of atelectasis, mucus plugging, pleural effusions, and parenchymal opacities that obscure the bronchial tree. This limitation is consistent with prior CF imaging work showing that severe structural abnormalities can reduce the reliability of automated segmentation and require more careful interpretation, particularly when the airway lumen is partially collapsed or surrounded by dense diseased parenchyma. In addition, because most CF CT images are acquired at full inspiration, methods developed for air-filled lungs may not generalize well to expiratory scans, where lung density is higher and regional air trapping becomes more prominent. Accordingly, additional validation on expiratory imaging would be needed to establish the framework’s robustness across acquisition states.

The framework described in this study can automatically generate airway trees and skeletonized representations, facilitating quantitative assessment of CF-related airway remodeling. Specifically, it can measure lung and airway volumes, branch counts, generational splits, airway diameters, and cross-sectional areas, all of which are established quantitative descriptors of airway morphology in CT-based CF studies. The stage-dependent increase in these metrics is also consistent with prior reports of progressive airway wall thickening, luminal narrowing due to mucus plugs, and broader structural airway abnormalities in children with CF over time. Taken together, these findings support the framework as a useful quantitative tool for characterizing CF airway disease, while highlighting the need for validation in more severe and expiratory imaging settings.

In summary, a major contribution of this study is the development of an automated, quantitative method for detecting and measuring airways in CT scans, enabling the assessment of CF severity based on changes in airway characteristics. However, as with all new analytical approaches, the current work has limitations. For instance, the framework can overestimate airways in severe cases, where conditions such as honeycomb patterns, clusters of bullae, and/or extensive atelectasis are present. Hence, this approach may be biased in severe cases, suggesting that airway analysis may be more suitable for detecting CF in early- to mild-stage cases. In addition, scanner resolution affects the accuracy of airway and volume measurements by limiting the smallest structures that can be measured. To overcome this limitation, the use of texture analysis to examine ultrastructural changes that may not be visually apparent has been reported to provide value. For example, texture has been used to distinguish between different tissue types and disease stages in cancer research by extracting texture features from medical images (CT, mammographic, and MRI). In the context of CF, we suspect that lung lesion patterns can characterize the disease. By quantifying these subtle variations in tissue texture, hidden patterns beyond resolution capabilities can be discovered.

Lastly, current clinical approaches for airway diseases rely on spirometry measurements, which may lack the sensitivity to detect subtle changes associated with disease progression or therapy response. Hence, to the best of our knowledge, there is no gold standard for quantitatively evaluating diseased airways and their impact on disease progression. Our approach is a first attempt to provide a sensitive framework for characterizing airways from a single CT volume.

## Acknowledgements

This work is supported by grants NIA U54 AG054345 and NIA R21 AG07857501.

## Author’s Contributions

JAKCC contributed to the study design, data collection and management, data analysis and interpretation, data quality assessment, literature review, and manuscript drafting. MLC participated in the study design, data collection, data analysis, and data interpretation. SAP and CPB helped in the data collection, analysis and quality assessment. PS provided statistical and image analysis advice, participated in data interpretation, and helped write the manuscript. PRT contributed to the study design, data analysis and interpretation, and helped write the manuscript. JAKCC, PRT, and SAC had access to all data. JAKCC, PS, and PRT verified the data and results. All authors helped revise the manuscript. All authors had final responsibility for the decision to submit for publication.

## Data Sharing

The diagnostic images analyzed in this retrospective study are the proprietary data of Indiana University Health (IU Health) and are subject to strict institutional data governance policies. To comply with patient privacy regulations and institutional restrictions, access to the minimal underlying dataset is restricted but may be made available to qualified researchers upon reasonable request, subject to formal approval by IU Health and the execution of a legally binding Data Transfer Agreement (DTA)

## Ethics Committee Approval

Ethics approval was not required for this study.

## Institutional Review Board Approval

The study was approved by the Indiana University Institutional Review Board (Protocol IU IRB #14345 Cystos - Artificial Intelligence Project).

