## Supplemental Figures for "Automated Airways Characterization and Assessment of Cystic Fibrosis from CT Imaging"

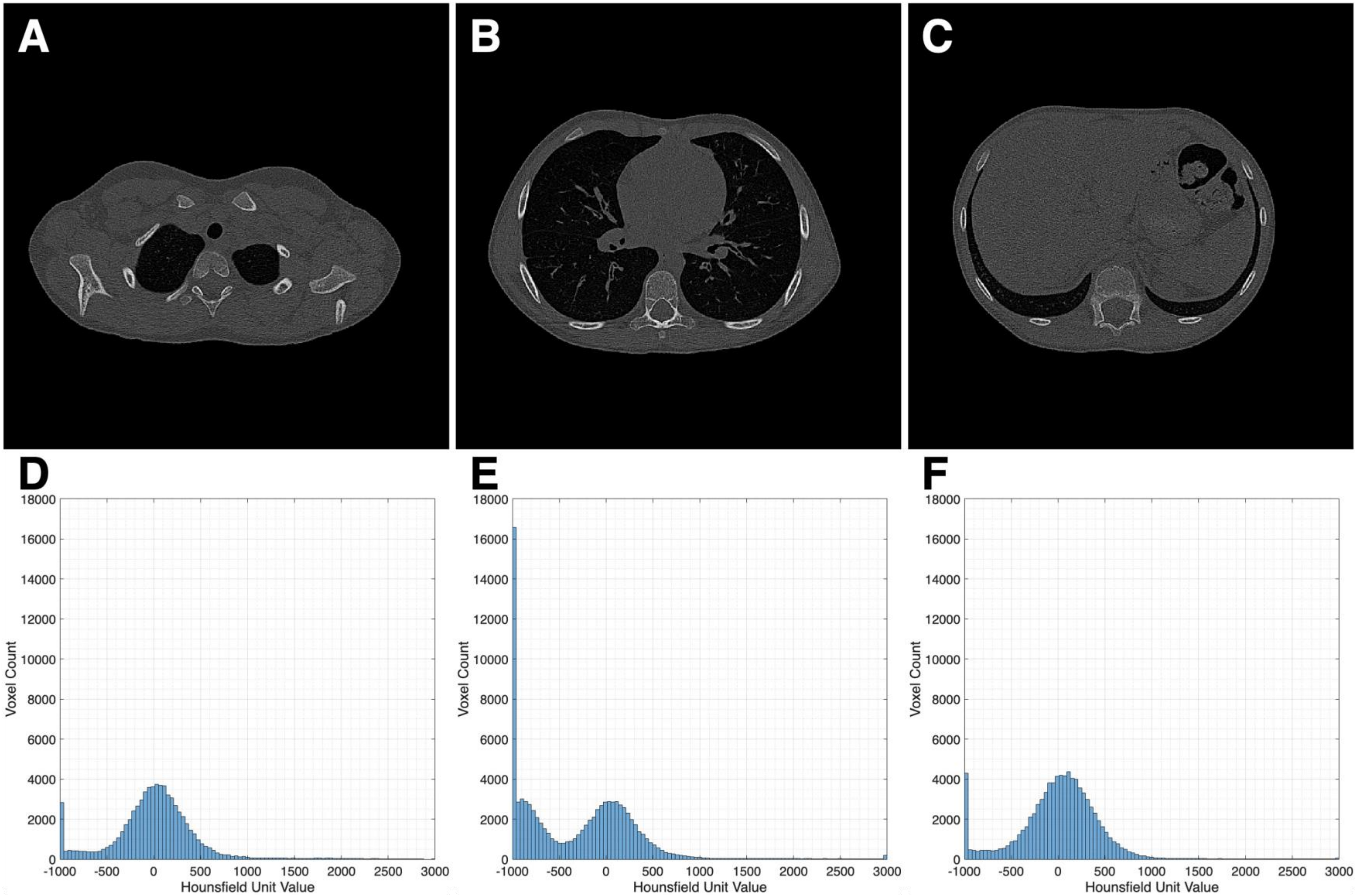


**Supplemental Figure S1 – Lung Shape at Different Locations**
Lung slices at (A) top, (B) middle, and (C) bottom, and their corresponding histograms for (D) top, (E) middle, and (F) bottom, which illustrate the different shapes of the lungs and histograms across different locations. For example, the top of the lung (A) appears quasi-circular, and it transitions into a thin "C" shape at the bottom (C). In addition, the peaks of the histograms vary in counts and in location.

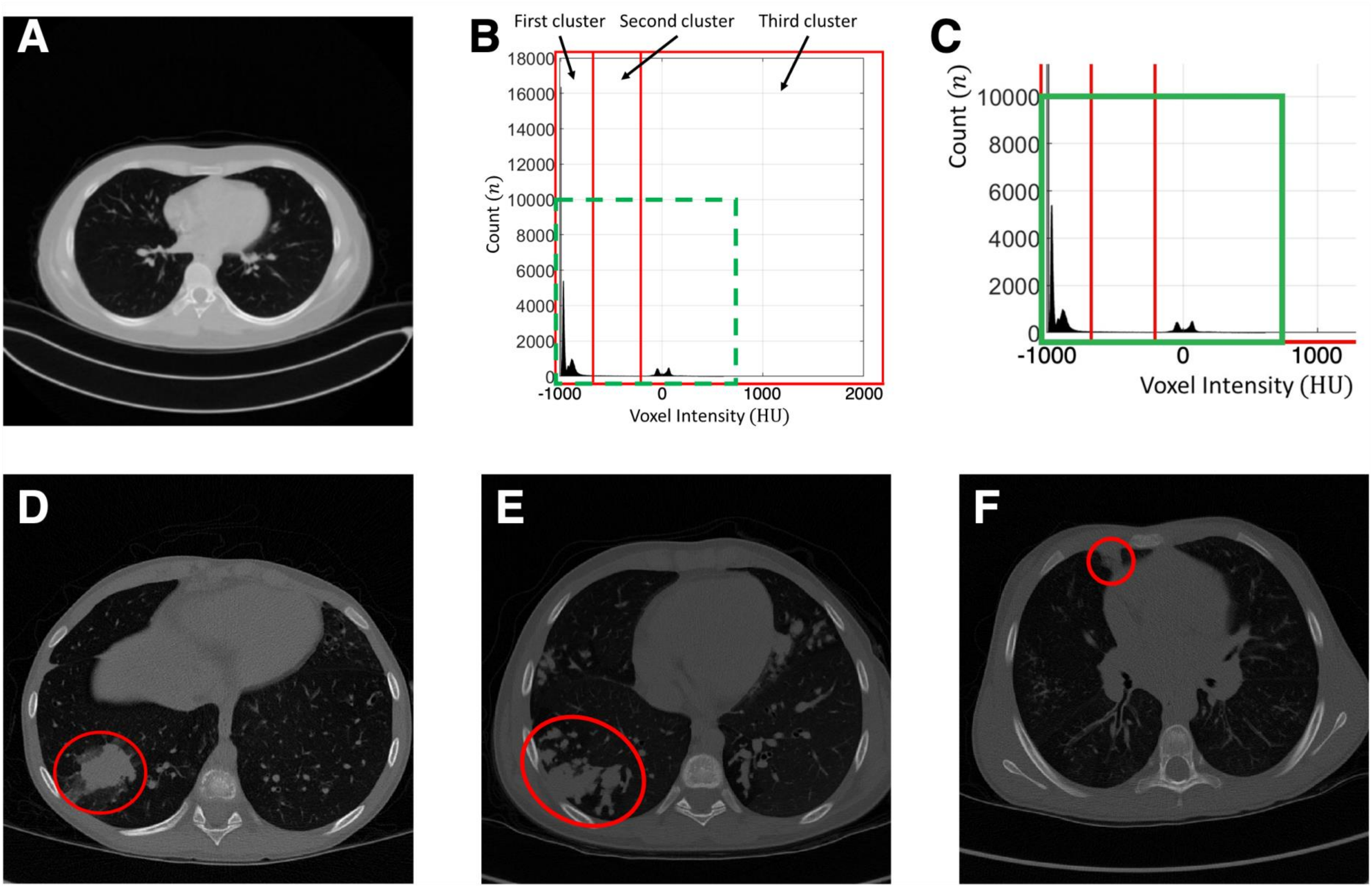


**Supplemental Figure S2 – Example of the 3-cluster Gaussian Mixture Model for a Singles Slice and Example of Abnormalities Observed in Severe Cases.**

(A) Single slice in the middle of the lung. (B) Histogram generated using the slice and its two consecutive slices (previous and next). (C) Inset of histogram in B. (D-F) Lung abnormalities observed in moderate to severe CF subjects.

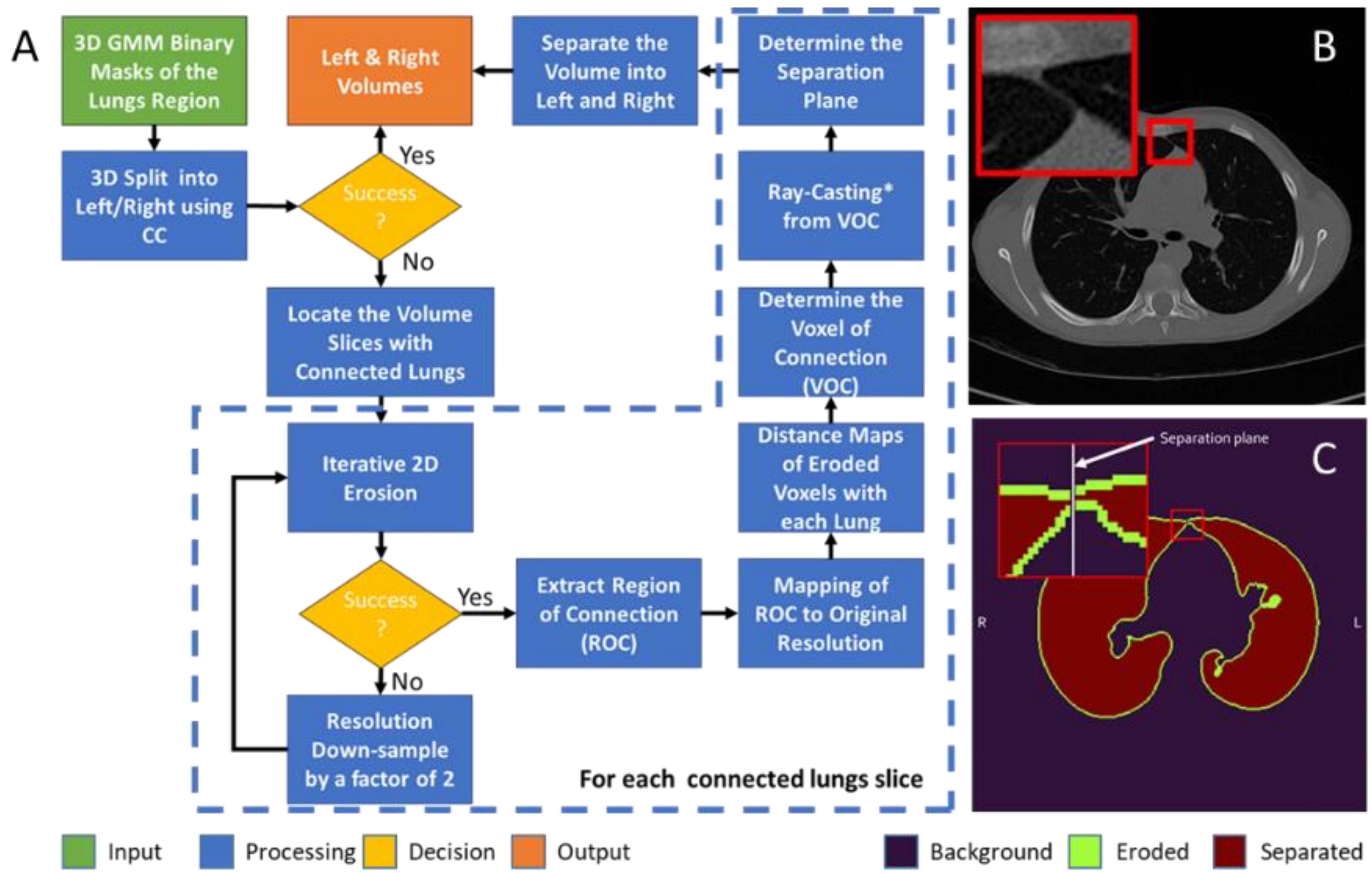


**Supplemental Figure S3 – SOLVR Algorithm**
(A) Workflow for the Separation of Lung Volume Regions (SOLVR). (B) CT Image with the inset illustrating the anterior sterno-pericardial ligaments, which prevent left-side and right-side separation. (C) An example of a lung mask illustrating the separation into left- and right-lung volumes using SOLVR.
